# Changes in psychiatric documentation and treatment in primary care with artificial intelligence scribe use

**DOI:** 10.1101/2025.05.14.25327620

**Authors:** Victor M. Castro, Thomas H. McCoy, Pilar Verhaak, Anudeepa K. Ramachandiran, Roy H. Perlis

**Affiliations:** Center for Quantitative Health and Department of Psychiatry, Massachusetts General Hospital, Boston, MA; Department of Psychiatry, Harvard Medical School, Boston, MA

**Author notes:** **Correspondence** Roy H. Perlis, Massachusetts General Hospital, 185 Cambridge Street, 6th Floor, Boston, MA 02114.

**Keywords:** Large language model, Artificial intelligence, Machine Learning, Major depressive disorder, Antidepressant, Research Domain Criteria

## Abstract

**Importance:** Despite increasingly widespread use of artificial intelligence-driven ambient scribes in medicine, the extent to which they may impact clinician practice is not well-studied.

**Objective:** To characterize differences in documentation and treatment of psychiatric symptoms in primary care outpatient notes generated using ambient scribes.

**Design:** Case-control electronic health records

**Setting:** Primary care annual visit notes from the Massachusetts General and Brigham and Women’s Hospital systems between February 2023 and February 2023.

**Participants:** Random sample of 20,302 notes from 4 types of visits, matching 1:1 using sociodemographic and clinical features: those using an ambient scribe, those using a human scribe, those occurring during the same period without a scribe, and those occurring prior to scribe deployment

**Exposure:** Use of an artificial intelligence-driven ambient scribe

**Main Outcome and Measures:** Neuropsychiatric symptom documentation, in terms of estimated Research Domain Criteria, using a HIPAA-compliant large language model (GPT4o; gpt-4o-11-20); incident antidepressant prescriptions and diagnostic codes; referral for mental health follow-up.

**Results:** In the ambient scribe group, mean age was 48 (SD 14) years; 59% of notes reflected individuals of female sex, and 5.0% met criteria for moderate or greater depressive symptoms by PHQ-9. Estimated levels of RDoC symptomatology in all 6 domains were significantly greater in the ambient-scribed notes (p <.001 for all contrasts). In a logistic regression model, likelihood of a psychiatric intervention (referral, new diagnosis, or antidepressant prescription) was significantly lower among ambient-scribed visits compared to unscribed (aOR 0.83, 95% CI 0.72-0.95), but not for human-scribed compared to unscribed (aOR 1.01, 95% CI 0.87-1.17).

**Conclusion and Relevance:** In this case-control design examining outpatient primary care notes, we found that incorporation of artificial intelligence-driven ambient scribes in primary care was associated with greater levels of neuropsychiatric symptom documentation but lesser likelihood of acting on psychiatric symptoms. Further study will be required to determine whether these changes are associated with differential outcomes.

**Trial registration:** n/a

**Key Points:** *Question:* How is documentation and treatment of psychiatric symptoms in primary care different for outpatient visits using artificial intelligence (AI)-driven ambient scribes.

*Findings:* In more than 20,000 routine annual visits, ambient scribe use was associated with greater documentation of neuropsychiatric symptoms but less likelihood of a depression-related intervention or diagnostic code.

*Meaning:* The extent to which use of ambient scribes may alter response to psychiatric symptoms by clinicians merits further investigation.

## Introduction

The use of artificial intelligence (AI)-driven ambient scribes, applying speech recognition and large language models to automate narrative note generation, has rapidly become widespread in medicine. To date, investigations of the impact of this transition are limited. One study suggested that clinicians using ambient scribes spent, on average, 5 minutes less per visit using the electronic health record^1^; others yielded mixed results^2,3,4^, with one study suggesting little change in measures of clinician productivity^5^. A broader set of studies have suggested that efforts to improve documentation in electronic health records correlate with improvements in overall quality of patient care.^6,7^

In particular, little is known about how the use of ambient scribes may change documentation and management of neuropsychiatric symptoms. Despite efforts to encourage more systematic symptom measurement for depression^8,9^, prior work suggested that incorporation of the PHQ-9 as a patient-reported outcome was associated with diminished documentation of depressive symptoms.^10^ It is possible that use of scribes could ameliorate this diminution, providing clinicians more time to discuss mental health with patients at annual visits and increasing the likelihood that such symptoms are documented. On the other hand, if mental health is not prioritized in the visit, no such shift would be observed. While prior findings could reflect a lack of documentation but not discussion, a lack of change here might indicate a lack of discussion.

To address this gap in knowledge, we drew on the electronic health records of outpatient primary care clinics of two large academic health systems, which span academic medical centers, community hospitals, and affiliated outpatient practices. To compare to notes generated via ambient scribing, we identified a contemporaneous group (i.e., from practices or clinicians not yet transitioned to ambient scribes), plus a second set of notes from a prior year to exclude the possibility that differences reflected enthusiasm for using this technology. We hypothesized that, with the shift to ambient scribes, documentation of neuropsychiatric symptomatology would be greater than without scribes.

## Methods

### Study design and cohort derivation

From the electronic health records of two large academic medical centers in Eastern Massachusetts, we drew a random sample of 5,076 outpatient annual visit notes, identified by a corresponding Current Procedural Terminology (CPT) code (99385-99387, 99395-99397, or HCPCS G0402, G0438, G0439), among individuals age 18 or older seen between February 2024 and February 2025 who had an associated PHQ score recorded, and whose note included documentation of ambient scribe use. Ambient scribe use was determined based on documentation of patient consent to use of the technology in the clinical note. For comparison, we drew 3 additional samples of outpatient annual visit notes matched 1:1 on age at visit, sex, race and prior depressive disorder diagnosis (International Classification of Diseases (ICD)-10 codes F06.3*, F32.*, F33.*, F34.1, and F53.0): one including documentation of a human scribe (identified using standard text in the clinical note), one including no such documentation between February 2024 and February 2025 (the contemporaneous sample), and one with no such documentation from a period before ambient scribe use, between February 2023 and January 2024.

A total of 20,302 notes across 4 types of visits were included in analysis. For each visit, we identified sociodemographic features include sex, age at visit, self-reported race and ethnicity, highest level of education, as well as insurance status (commercial or otherwise), in order to facilitate comparisons of the cohorts. Additionally, we identified ICD codes corresponding to the visit, RxNorm codes indicating the prescription of an antidepressant (including SSRIs, SNRIs, TCAs, bupropion, and trazodone), clinician orders for ambulatory referrals, and corresponding PHQ scores. The study was approved by the Institutional Review Board of Mass General-Brigham, granting a waiver of informed consent in accordance with 45 CFR 46.116 as it would be infeasible to contact participants and no participant interaction was required.

### Generation of estimated Research Domain Criteria scores

In prior work, we have demonstrated that language models can readily estimate dimensions of NIMH Research Domain Criteria (RDoC)^11^ symptomatology^12^. We applied this methodology to characterize such scores in each narrative clinical note. Specifically, we applied a Python script to present individual narrative clinical notes via an application programming interface (API) to a HIPAA-compliant instance of GPT4o (gpt-4o-11-20) hosted by Microsoft Azure, with model temperature set at 0 to yield results as deterministic as possible. The prompt was “You are a skilled psychiatrist scoring an outpatient visit note in terms of how the patient symptoms over the past 24 hours reflect the 6 RDoC domains: Negative Valence Systems, Positive Valence Systems, Cognitive Systems, Social Processes, Arousal and Regulatory Systems, and Sensorimotor Systems. Remember that substance use can be reflected as Positive Valence symptom. Notes are scored on a 0–10 scale to capture the magnitude of documented symptoms relevant in a given domain. Score 0 if no symptoms are present and functioning is normal, 1–3 mild symptoms, 4–6 moderate, 7–9 severe, 10 extremely severe. Generally, moderate requires treatment and severe requires hospitalization. Return only a JSON object with each score, Negative, Positive, Cognitive, Social, Arousal, and Sensorimotor.” The model was re-initialized after each presentation to ensure prior notes could not influence subsequent scores.

### Analysis

In univariate analyses, we compared features of ambient AI-scribed notes to the comparator groups using Mann-Whitney test or chi square test. We similarly compared estimated RDoC domain scores and estimated proportion of visit devoted to mental health, and to depression in particular. These analyses focused on comparing AI-scribed to human-scribed, and to contemporaneous unscribed notes; we also included a prior unscribed set (i.e., before AI scribes were deployed) to exclude the possibility that contemporaneous unscribed notes reflected other differences. We primarily examined an aggregate outcome of depression diagnosis, antidepressant prescription, or referral for psychiatric evaluation and care; we also examined each outcome individually. Beyond univariate contrasts, we fit multiple logistic regression models to examine associations between scribe type and likelihood of such follow-up, adjusted for sociodemographic features.

A nominal p-value of 0.05, two-tailed, was set as the threshold for statistical significance. Bonferroni correction for note length, 6 RDoC domains and 2 additional measures, as well as the composite outcome, would require p <.005 for statistical significance. All analyses used R 4.4.0^13^.

## Results

Characteristics of individuals reflected in the visit notes from each of the four groups are shown in Table 1. Mean age at visit was 48 years (SD 14) in all groups. Mean PHQ-9 scores were similar across groups: 1.2 (SD 3.5) for Ambient AI Scribe, 1.2 (SD 3.6) for Human Virtual Scribe, 1.2 (SD 3.5) for No Scribe, and 1.2 (3.6) for No Scribe (prior to ambient scribe technology deployment). The proportion of visits reflecting a PHQ-9 score greater than or equal to 10 was also similar: 254 (5.0%) for Ambient AI Scribe, 256 (5.0%) for Human Virtual Scribe, 265 (5.2%) for No Scribe (after deployment), and 251 (49%) for No Scribe (prior to deployment).

**Table 1.** Characteristics of individuals in the ambient scribe and comparator cohorts.

| Characteristic | Ambient AI Scribe<br>N = 5,076<sup>1</sup> | Human Virtual Scribe<br>N = 5,075<sup>1</sup> | No Scribe<br>(post deployment)<sup>2</sup><br>N = 5,075<sup>1</sup> | No Scribe<br>(prior to deployment)<sup>3</sup><br>N = 5,076<sup>1</sup> |
| --- | --- | --- | --- | --- |
| Age at visit (y) | 48 (14) | 48 (14) | 48 (14) | 48 (14) |
| Female sex | 2,989 (59%) | 3,002 (59%) | 2,981 (59%) | 2,988 (59%) |
| Race |  |  |  |  |
| Asian | 567 (11%) | 574 (11%) | 566 (11%) | 567 (11%) |
| Black | 344 (6.8%) | 320 (6.3%) | 351 (6.9%) | 318 (6.3%) |
| Other | 564 (11%) | 561 (11%) | 560 (11%) | 590 (12%) |
| White | 3,601 (71%) | 3,620 (71%) | 3,598 (71%) | 3,601 (71%) |
| Hispanic ethnicity | 519 (10%) | 576 (11%) | 500 (9.9%) | 527 (10%) |
| At least Bachelor's degree | 3,035 (60%) | 2,948 (58%) | 2,914 (57%) * | 2,851 (56%) ** |
| Public insurance | 250 (4.9%) | 172 (3.4%) *** | 199 (3.9%) * | 224 (4.4%) |
| Depression code in prior<br>2 years (ICD10) | 867 (17%) | 991 (20%) ** | 996 (20%) ** | 955 (19%) * |
| PHQ-9 score | 1.2 (3.5) | 1.2 (3.6) | 1.2 (3.5) | 1.2 (3.6) |
| PHQ-9 ≥ 10 | 254 (5.0%) | 256 (5.0%) | 265 (5.2%) | 251 (4.9%) |

<sup>1</sup> Mean (SD), \* Bonferroni corrected $p < 0.05$ , \*\* $p < 0.01$ , \*\*\* $p < 0.001$
<sup>2</sup> Notes written after the deployment of ambient scribe technology
<sup>3</sup> Notes written prior to the deployment of ambient scribe technology

We first compared overall note length between groups. Length was greatest in the Human Virtual Scribe group, with a mean of 16,252 characters (SD 7,060), followed by Ambient AI Scribe at 13,629 (SD 4,507), and No Scribe at 7,932 (SD 5,025); p <.001 for all contrasts. We then examined RDoC domain scores estimated from the note text. Across all six domains, scores were significantly higher in the Ambient AI Scribe group compared to the other groups (Figure 1 and Table 2). For Negative Valence, scores were 2.05 (SD 1.82) in Ambient AI Scribe, 1.79 (SD 1.77) in Human Virtual Scribe, 1.57 (SD 1.77) in No Scribe (after deployment), and 1.55 (SD 1.75) in No Scribe (prior to deployment). Positive Valence scores were 3.14 (SD 2.22), 2.74 (SD 2.19), 2.34 (SD 1.99), and 2.29 (SD 1.98) respectively. Cognitive domain scores were 1.66 (SD 1.48), 1.37 (SD 1.49), 1.27 (SD 1.48), and 1.26 (SD 1.49); Social domain: 1.70 (SD 1.70), 1.46 (SD 1.63), 1.28 (SD 1.57), and 1.26 (SD 1.57); Arousal: 2.84 (SD 1.83), 2.43 (SD 1.83), 2.05 (SD 1.85), and 2.03 (SD 1.83); Sensorimotor: 2.33 (SD 1.70), 1.77 (SD 1.62), 1.54 (SD 1.59), and 1.53 (SD = 1.60). All contrasts were statistically significant at p < 0.001.

**Table 2.** Comparison of note length and large language model-computed RDoC domain scores in note text.

| **Characteristic** | **Ambient AI Scribe**<br>N = 5,076<sup>1</sup> | **Human Virtual Scribe**<br>N = 5,075<sup>1</sup> | **No Scribe (post deployment)**<br>N = 5,075<sup>1</sup> | **No Scribe (prior to deployment)<sup>2</sup>**<br>N = 5,076<sup>1</sup> |
| --- | --- | --- | --- | --- |
| Note length (characters) | 13,629 (4,507) | 16,252 (7,060) *** | 7,932 (5,025) *** | 7,489 (4,738) *** |
| RDoC: Negative valence | 2.05 (1.82) | 1.79 (1.77) *** | 1.57 (1.77) *** | 1.55 (1.75) *** |
| RDoC: Positive valence | 3.14 (2.22) | 2.74 (2.19) *** | 2.34 (1.99) *** | 2.29 (1.98) *** |
| RDoC: Cognitive | 1.66 (1.48) | 1.37 (1.49) *** | 1.27 (1.48) *** | 1.26 (1.49) *** |
| RDoC: Social | 1.70 (1.70) | 1.46 (1.63) *** | 1.28 (1.57) *** | 1.26 (1.57) *** |
| RDoC: Arousal | 2.84 (1.83) | 2.43 (1.83) *** | 2.05 (1.85) *** | 2.03 (1.83) *** |
| RDoC: Sensorimotor | 2.33 (1.70) | 1.77 (1.62) *** | 1.54 (1.59) *** | 1.53 (1.60) *** |

<sup>1</sup>Mean (SD), \* Bonferroni corrected p< 0.05, \*\* p <0.01, \*\*\* p <0.001
<sup>2</sup> Notes written after the deployment of ambient scribe technology
<sup>3</sup> Notes written prior to the deployment of ambient scribe technology

**Figure 1.**
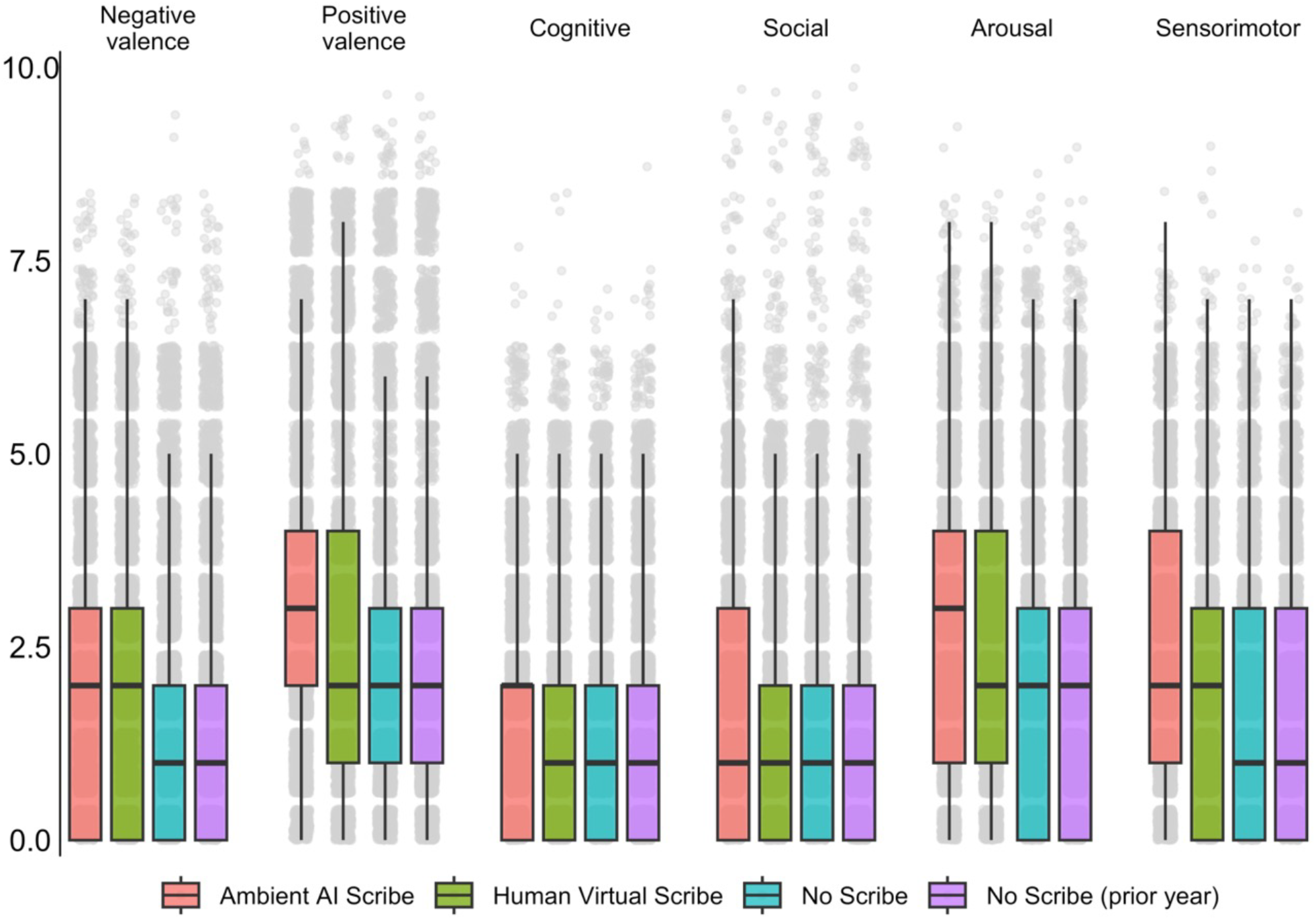
Estimates of RDoC symptomatology, by visit type

Visits were associated with a depression-related ICD-10 code in 447 (8.8%) of Ambient AI Scribe visits, 587 (12%) of Human Virtual Scribe visits, 604 (12%) of No Scribe (post deployment) visits, and 535 (11%) of No Scribe (prior to deployment) visits; p<.001 for the Ambient AI Scribe compared to each of the other groups (Table 3). Antidepressants were prescribed in 275 (5.4%), 313 (6.2%), 317 (6.2%), and 319 (6.3%) of visits, respectively; none of these contrasts were statistically significant. Similarly, a behavioral health referral was entered in 151 (3.0%), 133 (2.6%), 150 (3.0%), and 113 (2.2%) of visits, differences which were not statistically significant. Finally, the composite depression outcome—defined as presence of any of diagnosis, antidepressant prescription, or behavioral health referral—was lower in the Ambient AI Scribe group at 708 (14%) compared to 843 (17%) in the Human Virtual Scribe group, 855 (17%) in the No Scribe (post deployment) group, and 805 (16%) in the No Scribe (prior to deployment group); p<.001 for contrasts to the Ambient AI Scribe group.

**Table 3.** Comparison of depression interventions at annual visit.

| **Characteristic** | **Ambient AI Scribe N = 5,076<sup>1</sup>** | **Human Virtual Scribe N = 5,075<sup>1</sup>** | **No Scribe (post deployment)<sup>2</sup> N = 5,075<sup>1</sup>** | **No Scribe (prior to deployment)<sup>3</sup> N = 5,076<sup>1</sup>** |
| --- | --- | --- | --- | --- |
| Depression code at visit (ICD10) | 447 (8.8%) | 587 (12%) *** | 604 (12%) *** | 535 (11%) ** |
| Antidepressant prescription | 275 (5.4%) | 313 (6.2%) | 317 (6.2%) | 319 (6.3%) |
| Incident antidepressant prescription | 52 (1.0%) | 50 (1.0%) | 66 (1.3%) | 88 (1.7%) ** |
| Referral for behavioral health (order) | 151 (3.0%) | 133 (2.6%) | 150 (3.0%) | 113 (2.2%) |
| Composite depression outcome | 708 (14%) | 843 (17%) *** | 855 (17%) *** | 805 (16%) * |

<sup>1</sup>N(%), \* Bonferroni corrected $p < 0.05$ , \*\* $p < 0.01$ , \*\*\* $p < 0.001$
<sup>2</sup> Notes written after the deployment of ambient scribe technology
<sup>3</sup> Notes written prior to the deployment of ambient scribe technology

While the groups were matched on age, sex, and race and ethnicity, we next examined whether scribe use was associated with these differential outcomes after accounting for sociodemographic and clinical features. In a logistic regression model (Figure 2), likelihood of any psychiatric intervention was significantly lower among AI-scribed visits compared to unscribed (aOR 0.83, 95% CI 0.72-0.99); no difference between human-scribed (aOR 0.97, 95% 0.85-1.11).and unscribed visits (prior to deployment) (0.94, 95% 0.82-1.07) was observed.

**Figure 2.**
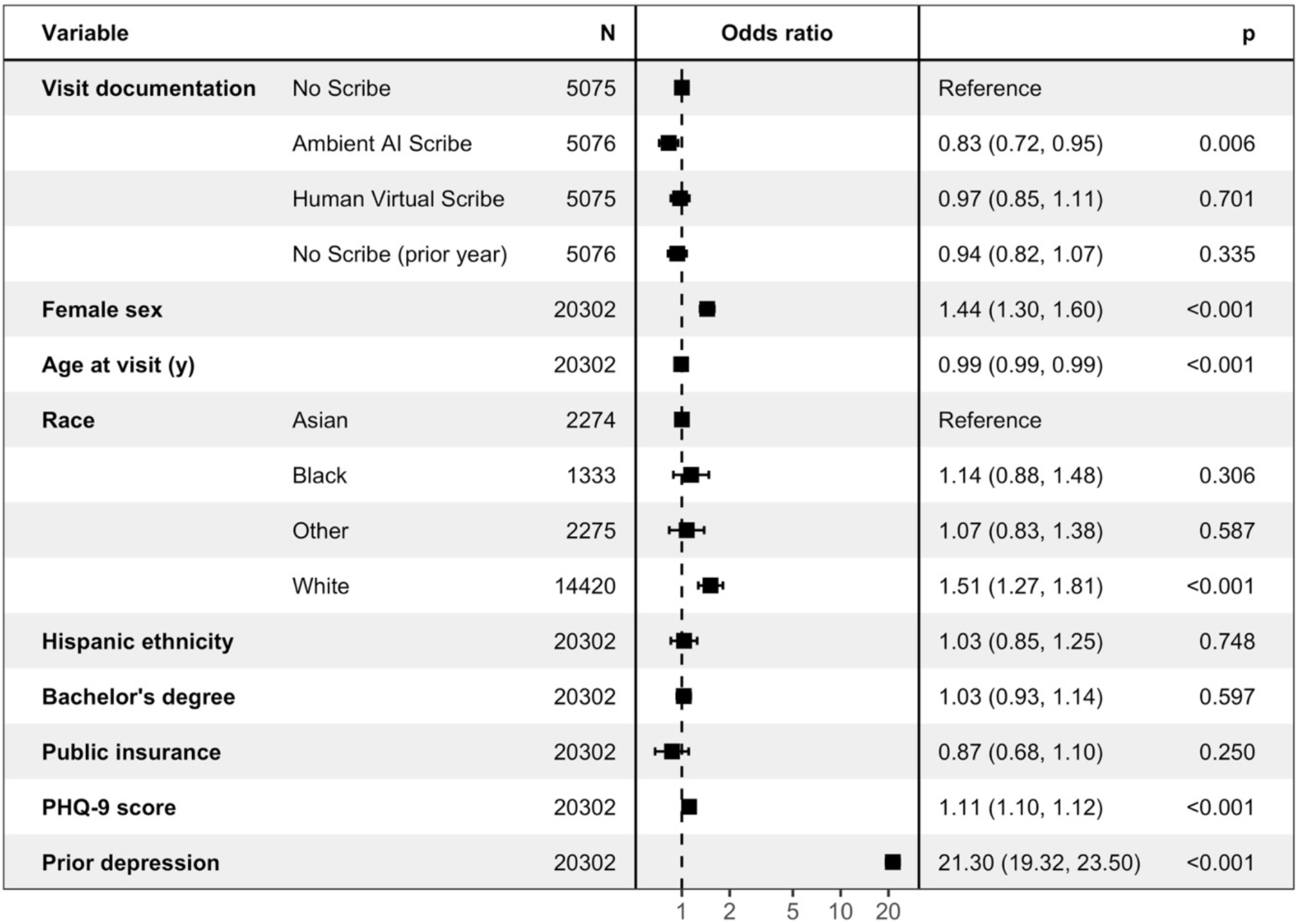
Logistic regression model of psychiatric intervention at visit

## Discussion

In this study examining clinical documentation from more than 20,000 outpatient annual visits, including roughly 5,000 incorporating AI scribes, we found that use of these scribes was associated with greater documented levels of neuropsychiatric symptoms compared to human scribes or no scribe conditions, but lesser likelihood of a depression intervention.

Some^2,3,4^ but not all^5^ studies have suggested that ambient scribes may reduce clinician workload, with concomitant improvement in clinician satisfaction^3,14,4^, but we could not identify comparable studies examining documentation itself. Other efforts to structure and standardize clinical documentation have generally been associated with improved note quality^6,7^. Conversely, one randomized trial of templated notes found that, while organization of such notes improved, they were felt to be less useful and to have lower accuracy when evaluated by clinical faculty^15^. More generally, the potential adverse impacts of language models on the reliability of clinical documentation was suggested by a recent commentary^16^. In this regard, our results are reassuring, suggesting that AI scribes in primary care have the potential to increase documentation of neuropsychiatric symptoms.

On the other hand, this increased documentation did not translate to increased attention to depressive symptoms: visits incorporating AI scribes were actually less likely to be associated with psychiatric interventions. One explanation for this association could be that automating documentation leads clinicians to be less active in general, analogous to reduced proficiency observed in pilots after the emergence of autopilot^17^. While this effect was not observed in human-scribed visits, it is possible that clinician attentiveness varies depending on the nature of the scribe.

In general, the rapid dissemination of AI scribes in medicine poses both an opportunity and a risk. Our results hint at the opportunity, if greater documentation of symptoms equates to greater response to those symptoms. On the other hand, many interventions in medicine have been adopted without clear evidence of benefit – particularly those, like scribes, that do not require formal regulatory review to establish effectiveness. To date, most investigations of AI scribes focus on impact on providers, but less so on patient care. Prospective studies will be valuable in understanding the potential for scribes to improve care – but also to understand their liabilities. Our work suggests scribes do change documentation substantially, but the impact on care quality remains to be determined.

### Limitations

This study has multiple limitations. While we generated two different types of comparison groups – one preceding deployment of ambient scribes, the other contemporaneous with use of scribes – we cannot exclude the possibility that associations between scribe use and measured note characteristics are confounded in other ways. The rapid dissemination of scribes may preclude randomized trials, but at minimum larger-scale prospective studies may be needed to determine causal effects. For example, differences in patient populations not captured by sociodemographic features could account for differential documentation. We also note that all notes reflect the use of ambient scribes in a single geographic region among affiliated health systems, albeit in varied clinical settings. Whether these effects apply in other populations, or generalize to all ambient scribe systems, remains to be investigated.

### Conclusions

While ambient scribes have rapidly become a standard approach to clinical documentation, this analysis of over 20,000 outpatient annual visit notes suggests that visits employing AI scribes may differ in clinically meaningful ways from those that rely on human scribes or do not use scribes at all. Our work suggests that further investigation will be critical to understand the potential for AI scribe use, and application of language models for documentation more generally, to impact neuropsychiatric symptom documentation in primary care and understand potential consequences.

## Data Availability

Due to the sensitive nature of EHR data, we are unable to make available the data used in the present work.

## Author Contributions

RHP and VMC conceptualized the study, conducted formal analyses, and drafted the manuscript. AKR and PFV helped draft and review the manuscript.

## Access to Data

Dr. Perlis had full access to all the data in the study and takes responsibility for the integrity of the data and the accuracy of the data analysis.

